# Influenza Vaccination and Disease Severity Among Older Adults Hospitalized with Influenza in France: A Nationwide Cohort Study, 2016-2024

**DOI:** 10.64898/2026.11.10.26367727

**Authors:** Claire Nour Abou Chakra, Aby Ba Diallo, André Some, Baptiste Pitel, Hélène Bricout, Clotilde El Guerche-Seblain, Cécile Chauvel, Marta C. Nunes

## Abstract

**Objective:** Seasonal influenza remains a major cause of severe morbidity and mortality among older adults, despite vaccine availability. We assessed whether influenza vaccination was associated with reduced disease severity among adults aged ≥65 years hospitalized with influenza in France.

**Methods and Analysis:** We conducted a nationwide retrospective cohort study using the French National Health Data System (SNDS). Adults aged ≥65 years hospitalized with confirmed influenza (ICD-10 codes: J09 or J10) between October 2016 and December 2024 were included, excluding 2020–2021. Vaccination status was identified from reimbursed community pharmacy dispensing records. Entropy balancing was used to balance sociodemographic characteristics, chronic conditions pneumococcal vaccination, and COVID-19 vaccination. Weighted regression models estimated adjusted risk ratios (aRRs) for invasive mechanical ventilation (IMV), intensive care unit (ICU) admission, in-hospital death, and a composite severe outcome. Multistate survival models assessed transitions specific hazard ratios (HR) between hospitalization, ICU, discharge, and in-hospital death.

**Results:** Among 132,443 influenza-associated hospitalizations, 72,852 (55%) occurred among vaccinated patients and 59,591 (45%) among unvaccinated patients. Vaccinated patients were older (mean age, 82.3 vs 79.3 years). IMV occurred in 2128 vaccinated (2.9%) and 2544 unvaccinated (4.3%) hospitalizations (aRR, 0.88; 95%CI: 0.83 to 0.93). In-hospital death occurred in 6300 (8.6%) and 4952 (8.3%), respectively (aRR, 0.90; 95%CI: 0.87 to 0.93). Vaccination was also associated with a lower risk of the composite outcome (aRR, 0.96; 95%CI: 0.94 to 0.98), but not ICU admission (aRR, 0.99; 95%CI: 0.96 to 1.01). Among patients admitted to ICU, vaccination was associated with a lower hazard of death (adjusted hazard ratio, 0.85; 95%CI: 0.78 to 0.92) and a 2-day reduction in the median time to discharge.

**Conclusion:** In this nationwide cohort study, influenza vaccination was associated with lower risks of IMV and in-hospital death among older adults hospitalized with influenza. These findings are consistent with possible attenuation of disease severity and support efforts to improve influenza vaccination among older adults.

## INTRODUCTION

Seasonal influenza remains a major cause of morbidity, hospitalization and mortality worldwide despite the long-standing availability of effective vaccines and established prevention strategies. Globally, influenza epidemics are estimated to cause approximately one billion infections each year, including 3-5 million cases of severe illness and between 290,000 and 650,000 respiratory deaths annually^1–3^. Older adults bear a disproportionate share of this burden owing to immunosenescence, multimorbidity, and increased susceptibility to influenza-associated complications^4,5^. In Europe, adults aged ≥65 years represent up to half of influenza-related hospitalizations and approximately 90% of influenza-associated deaths^4^.

In France, seasonal influenza epidemics represent a substantial burden on healthcare services, resulting on average in more than one million primary care consultations, over 20,000 hospitalizations, and approximately 9000 influenza-associated deaths each season, with the vast majority of severe outcomes occurring among older adults^6^. Influenza vaccination is recommended annually for individuals at increased risk of complications, including adults aged ≥65 years, individuals with chronic medical conditions, pregnant women, and healthcare workers. Despite these recommendations and full reimbursement for eligible groups, influenza vaccine coverage in France remains suboptimal, typically ranging between approximately 45% and 60% among adults aged ≥65 years^7^.

Although influenza vaccine effectiveness is traditionally evaluated against laboratory-confirmed infection or influenza-associated hospitalization, increasing attention has focused on whether vaccination also attenuates disease severity among patients who experience breakthrough influenza infection. Observational studies have reported that vaccinated patients hospitalized with influenza may experience reduced risks of severe outcomes, including intensive care admission and death^8–11^. However, the magnitude, consistency, and generalizability of this effect on disease severity remain uncertain. A narrative review highlighted substantial heterogeneity in the definition of severe outcomes and limited availability of large studies evaluating clinical severity among hospitalized patients with influenza^12^.

Nationwide administrative healthcare databases offer an opportunity to address these knowledge gaps by evaluating disease severity in routine clinical practice at the population level. In France, the national health data system (*Système National des Données de Santé*, SNDS) captures healthcare utilization and hospitalizations for nearly the entire French population^13^. Previous studies have used SNDS to evaluate influenza vaccination coverage, disparities in vaccine uptake, and the comparative effectiveness of different influenza vaccine formulations, including a recent nationwide analysis showing lower influenza-associated hospitalization rates among recipients of high-dose versus standard-dose quadrivalent vaccines^14–16^. These studies highlight the potential of SNDS data to investigate influenza vaccination outcomes in real-world populations.

Building on this nationwide data source, we aimed, to assess the severity of influenza-associated hospitalizations among vaccinated and unvaccinated older adults in France and to evaluate potential disease-attenuation effects of influenza vaccination across different clinically relevant outcomes.

## METHODS

We conducted a retrospective nationwide cohort study of adults aged ≥65 years hospitalized with influenza in France between 1 October 2016 and 31 December 2024 using data from SNDS. The SNDS includes anonymized individual-level information for more than 98% of the French population on outpatient healthcare utilization, hospital admissions (private and public facilities), reimbursed medications, and mortality ^13,17^. Individual healthcare encounters are linkable through unique anonymized patient identifiers.

The study followed the Strengthening the Reporting of Observational Studies in Epidemiology (STROBE) reporting guidelines.

### Study population

The study population was identified from the national hospital discharge database (*Programme de médicalisation des systèmes d’information-Médecine, Chirurgie, Obstétrique* [PMSI-MCO]), which includes discharge summaries of all admissions to public and private hospitals in France. Diagnoses are coded according to the International Classification of Diseases, 10^th^ revision (ICD-10), and procedures according to the French procedures’ classification (*Classification Commune des Actes Médicaux* [CCAM]). We identified all hospital admissions with an ICD-10 diagnosis code for influenza, including confirmed influenza with identified virus (J09 or J10) and influenza without virological identification (J11) (Supplementary Table-S1). Influenza codes could be recorded as primary, related, or associated diagnoses. These codes were extracted from discharge summaries and each medical unit summary in which patients were admitted during their hospitals stays.

For each individual patient, the index admission was defined as the first influenza-associated hospitalization during each epidemic season. Consecutive hospital stays separated by ≤1 day were combined and considered a single hospitalization episode.

Patients were excluded if they: i) were hospitalized in overseas French territories; ii) were admitted before the annual influenza vaccination campaign started; iii) received influenza vaccination <15 days before hospital admission; iv) received more than one influenza vaccine during the same season; v) resided in nursing homes with internal pharmacies in which individual vaccine dispensing is not captured in outpatient reimbursement databases; or vi) were hospitalized during the 2020–2021 season, which was heavily disrupted by the COVID-19 pandemic and unusually low influenza circulation. Influenza seasons were defined from 1 October of a given year to 30 September of the following year. Data for the 2024-2025 season were available until 31 December 2024 only.

### Influenza vaccination

Influenza vaccination status was identified using reimbursed outpatient pharmacy dispensing records from the DCIR database (*Datamart de Consommation Inter-Régimes*). In France, influenza vaccination is fully reimbursed for eligible high-risk groups, including adults aged ≥65 years, through a national voucher system allowing vaccine collection at community pharmacies. The date of pharmacy dispensing was used as a proxy for vaccination date. Influenza vaccine formulations were identified using Anatomical Therapeutic Chemical (ATC) classification codes and French drug identification codes (CIP13) (Supplementary Table-S2). The exposure to influenza vaccine was defined for each individual for every influenza season. Patients without a reimbursed influenza vaccine dispensing during a given season were classified as unvaccinated. Using the national surveillance data^18^, we also collected information on vaccine composition and the circulating influenza strains during each study season (Supplementary Table-S3).

### Outcomes and covariates

The main outcomes assessed during the index admission and compared between vaccinated and unvaccinated patients were: i) need for invasive mechanical ventilation (IMV); ii) admission to intensive care unit (ICU), defined as admission to an intermediate intensive care unit or reanimation unit; iii) in-hospital all-cause death; iv) composite severity outcome including IMV, ICU admission, or in-hospital death. Other indicators of disease severity among influenza-associated hospitalizations included length of hospital stay ≥5 days in those discharged alive, non-invasive mechanical ventilation, passive oxygen therapy, admissions to intermediate care or reanimation units, and influenza-related death identified using ICD-10 influenza codes (J09, J10 or J11) listed as the medical cause of death in the national death registry (*Centre d’épidémiologie sur les causes médicales de décès* [CépiDC]) available until 31 December 2023^17^. Outcomes at 30-days post-discharge among patients discharged alive, included rehospitalization for cardiorespiratory causes and all-cause death.

For each index admission the following variables were reported: i) Sociodemographic characteristics: age, sex, classification of the department of residence according to national data (urban, rural, or intermediate)^19^, residential setting (community dwelling or nursing home/social residence), and level of social deprivation measured using the French Deprivation Index (FDep) quintiles^20^; ii) Underlying chronic conditions were identified using a validated algorithm developed by the French National Health Insurance to identify 58 chronic health conditions among healthcare users (*Cartographie des pathologies et des dépenses*)^21^. Additional chronic conditions not fully captured by this algorithm were identified from PMSI diagnostic ICD-10 codes recorded during hospitalization. Chronic conditions were described individually and the number of chronic conditions per patient was categorized as none, one, two, or ≥3; iii) Acute cardiorespiratory, cerebrovascular, and renal events (Supplementary Table-S1) identified during hospitalization among patients without documented prior history of the corresponding chronic condition. Additionally, co-diagnoses of bacterial or other viral respiratory infections during admission; iv) Purchase of influenza antiviral treatment (oseltamivir or zanamivir) within 14-days before hospital admission, pneumococcal vaccination within the previous 5 years, and COVID-19 vaccination within the previous 12 months were identified using outpatient reimbursement records and ATC/CIP13 codes (Supplementary Table-S2).

### Statistical analyses

The data management was conducted within the secure SNDS environment using SAS software (SAS Institute Inc. 2023. SAS/STAT® 15.3. Cary, NC, USA) and statistical analyses were performed using R Statistical Software (v4.4.3; R Core Team 2021). The main analyses were restricted to hospitalizations with influenza ICD-10 codes J09 or J10 in any diagnostic position.

Baseline characteristics were compared between vaccinated and unvaccinated patients using standard descriptive statistics. Missingness was very low overall (<1%), except for FDep, that had approximately 8% missing data. Missing FDep values were imputed using multiple imputation by chained equations (MICE) based on patients’ baseline characteristics^22^. Complete-case analyses were performed for all other variables.

Because vaccinated and unvaccinated patients differed substantially in demographic and clinical characteristics, propensity score-based balancing approaches were used to reduce confounding. Two weighting methods were evaluated: Entropy Balancing (EBAL) and Inverse Probability of Treatment Weighting (IPTW). EBAL weights were obtained by solving an optimisation problem in which, for each covariate, the means and variances (in continuous cases) or the proportions (in discrete cases) in the vaccinated and unvaccinated patient groups are equal^23,24^. IPTW weights were estimated from the predicted probability of vaccination estimated through logistic regression^25^. Balancing models included epidemic season, age group (65-74, 75-84, ≥85 years), sex, classification of departments of residence, residential setting, FDep quintile, number of chronic conditions, pneumococcal and COVID-19 vaccination status. The performance of IPTW and EBAL was evaluated using standardized mean differences (SMD), with values <0.1 considered indicative of acceptable balance, maximum residual SMD, and overall SMD distribution (density plots).

To address temporal confounding introduced by the COVID-19 pandemic and availability of COVID-19 vaccines, three analytical scenarios were explored: i) separate analyses before and after COVID-19 vaccine availability; ii) inclusion of COVID-19 vaccination status with an interaction term for epidemic season (2021-2024); and iii) analyses without explicit accounting for COVID-19 vaccination. The final analytical strategy was selected based on covariate balance, weight stability, and consistency of effect estimates.

Associations between influenza vaccination and study outcomes were estimated by weighted logistic regression models and marginal method to derive risk ratios (RR), with 95% confidence intervals (CI) computed by the delta method^26^. Adjusted models additionally included antiviral treatment before admission, co-diagnosis of bacterial or viral infection, acute events identified during hospitalization, and week of admission. Week of admission was modelled using natural cubic splines with 5 knots to account for seasonal variation in influenza circulation and timing of vaccination. The knots were defined as weeks 7, 13, 18, 22 and 31 from the week of October 1^st^ of each season (calendar week 39 or 40 depending on the season). Subgroup analyses were conducted according to age group and epidemic season.

To characterize clinical trajectories following hospitalization, we performed multistate survival analyses within 30-days of admission using EBAL-derived weights. ICU admission was modelled as a transient state, whereas discharge alive and in-hospital death were treated as absorbing states. Adjusted Hazard ratios (aHR) and 95%CIs were estimated using weighted Cox models including the same covariates as the primary outcome analyses. State occupation probabilities and 95%CI were estimated using Aalen–Johansen estimator^27^.

Five sensitivity analyses were conducted to assess the robustness of findings: i) repeating the adjusted analyses without inclusion of acute events identified during hospitalization; ii) restricting analyses to hospitalizations with ICD-10 codes J09 and J10 recorded as the primary diagnosis; iii) including all influenza-associated hospitalizations, including J11 codes; iv) excluding hospitalizations with documented COVID-19 co-infection; v) restricting analyses to hospitalizations with pneumonia (ICD-10 J12-J18) recorded as the primary diagnosis.

### Ethics approval

According to French regulations (Public Health Code Articles L.1461–3 and R.1461–11), research using pseudonymized data from the SNDS does not require ethics committee review or individual informed consent. Conducting the study did not require specific authorization form the French data protection authority. The study has been declared prior to data extraction on the Health Data Hub online platform (No. 24832545).

### Patient and public involvement

Patients and members of the public were not involved in the design, conduct, analysis, or reporting of this study, which used routinely collected, pseudonymized national healthcare data.

### Dissemination of results

Preliminary findings from this study were presented at the Options for the Control of Influenza XIII conference in 2026. Following publication, the results will be disseminated through the CERP website and social media channels to reach healthcare professionals, researchers, and the wider public. Individual participants will not be contacted because the study used pseudonymized national healthcare data without direct participant recruitment.

## RESULTS

### Study population

Between 1 October 2016 and 31 December 2024, 171,875 hospital admissions with an influenza diagnosis were identified among adults aged ≥65 years (Supplementary Figure-S1). After applying exclusion criteria, 149,210 index hospitalizations remained, including 132,443 with virologically confirmed influenza (ICD-10 J09 or J10), of which 72,852 (55.0%) occurred among vaccinated patients and 59,591 (45.0%) among unvaccinated patients. Among confirmed influenza admissions, influenza was recorded as primary diagnosis in 62.7% (n=82,798) of them (Supplementary Table-S4). Weekly influenza-associated hospitalizations trends were broadly similar between vaccinated and unvaccinated patients across epidemic seasons (Supplementary Figure-S2).

Before weighting, vaccinated patients were older than unvaccinated patients (mean age 82.3 vs 79.3 years) and more frequently had underlying chronic conditions (92.4% vs 85.9%), pneumococcal vaccination (16.6% vs 7.9%), and COVID-19 vaccination (62.2% vs 28.5%) (Supplementary Table-S4). Differences were also observed according to epidemic season, FDep, residential setting, and area of residence.

Among vaccinated patients, the mean interval between influenza vaccination and hospital admission was 92.8 days and more than 70% had been vaccinated >60 days before admission (Supplementary Table-S4). The most commonly administered vaccines were standard-dose injectable formulations (92.5%), whereas high-dose accounted for a minority of vaccinations (7.5%).

Both IPTW and EBAL substantially improved covariate balance, with SMD of ≤0.1 for all balancing variables (Table-1), with EBAL achieving a slightly better overlap in weighted propensity score distributions than IPTW (Supplementary Figures-S3, S4).

**Table 1.**
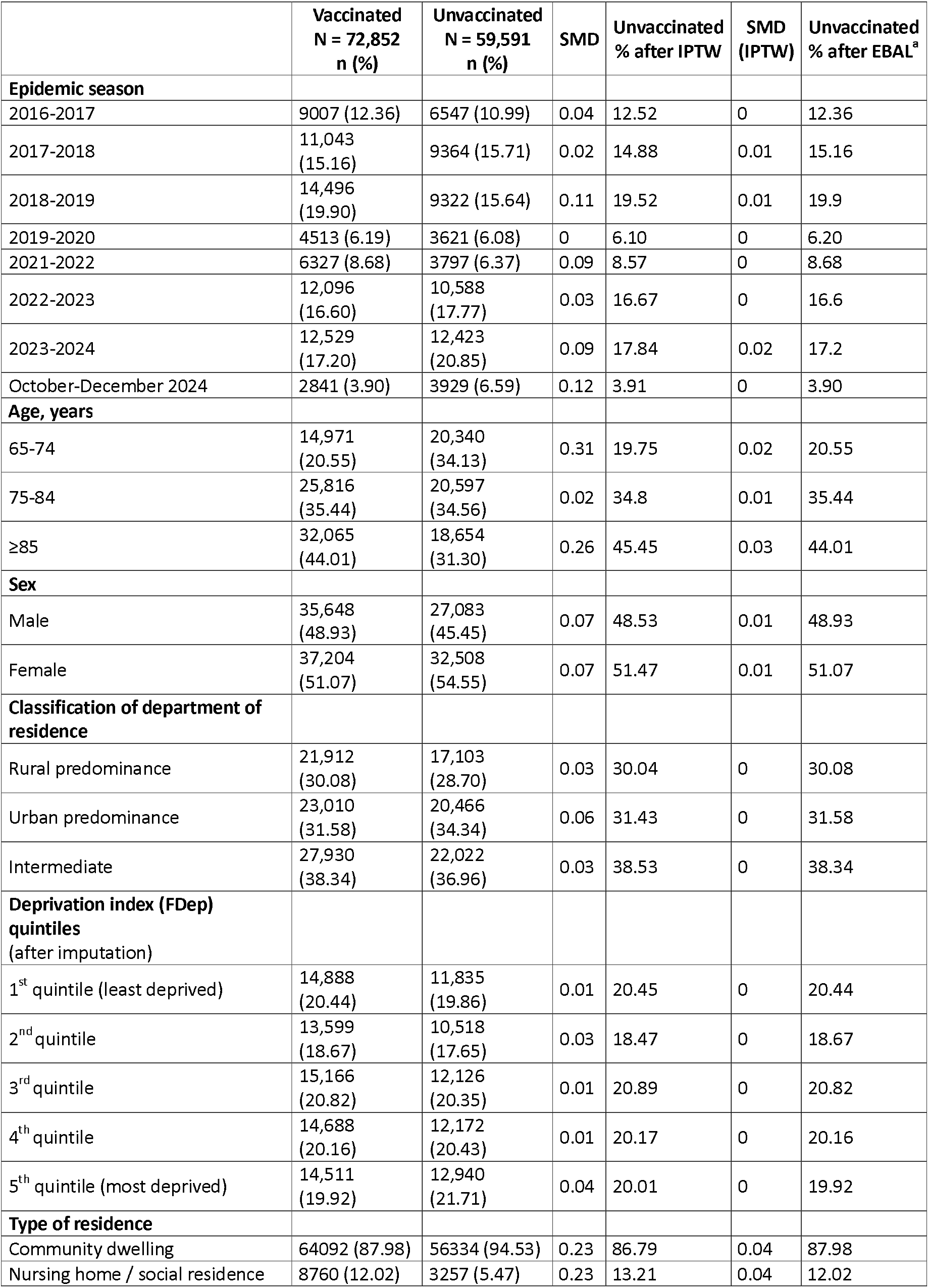
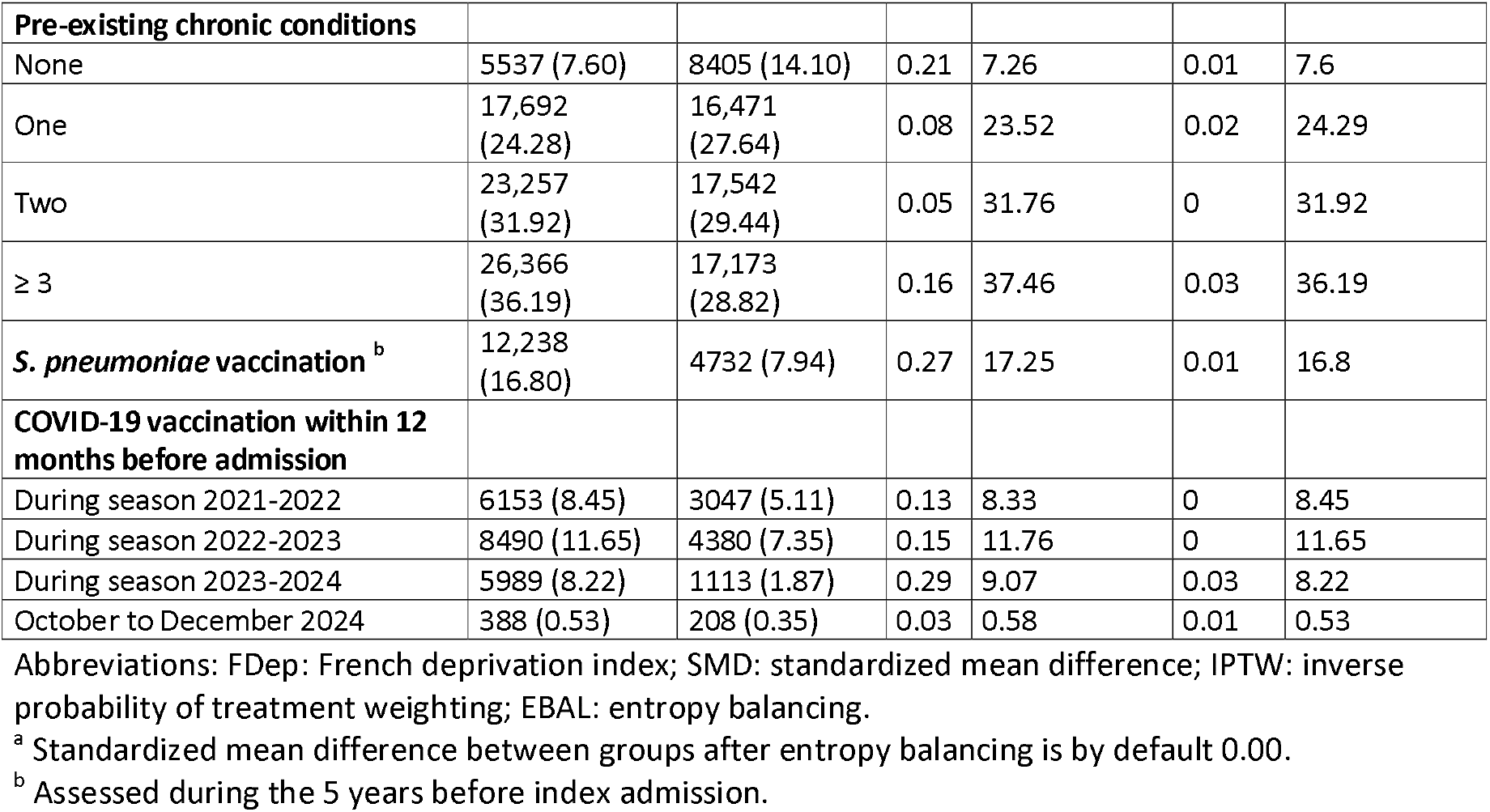
Distribution of patients’ characteristics among vaccinated and unvaccinated patients hospitalized with a diagnosis of influenza confirmed infection before balancing and after inverse probability of treatment weighting or entropy balancing.

|  | Vaccinated<br>N = 72,852<br>n (%) | Unvaccinated<br>N = 59,591<br>n (%) | SMD | Unvaccinated<br>% after IPTW | SMD<br>(IPTW) | Unvaccinated<br>% after EBAL<sup>a</sup> |
|---|---|---|---|---|---|---|
| **Epidemic season** |  |  |  |  |  |  |
| 2016-2017 | 9007 (12.36) | 6547 (10.99) | 0.04 | 12.52 | 0 | 12.36 |
| 2017-2018 | 11,043 (15.16) | 9364 (15.71) | 0.02 | 14.88 | 0.01 | 15.16 |
| 2018-2019 | 14,496 (19.90) | 9322 (15.64) | 0.11 | 19.52 | 0.01 | 19.9 |
| 2019-2020 | 4513 (6.19) | 3621 (6.08) | 0 | 6.10 | 0 | 6.20 |
| 2021-2022 | 6327 (8.68) | 3797 (6.37) | 0.09 | 8.57 | 0 | 8.68 |
| 2022-2023 | 12,096 (16.60) | 10,588 (17.77) | 0.03 | 16.67 | 0 | 16.6 |
| 2023-2024 | 12,529 (17.20) | 12,423 (20.85) | 0.09 | 17.84 | 0.02 | 17.2 |
| October-December 2024 | 2841 (3.90) | 3929 (6.59) | 0.12 | 3.91 | 0 | 3.90 |
| **Age, years** |  |  |  |  |  |  |
| 65-74 | 14,971 (20.55) | 20,340 (34.13) | 0.31 | 19.75 | 0.02 | 20.55 |
| 75-84 | 25,816 (35.44) | 20,597 (34.56) | 0.02 | 34.8 | 0.01 | 35.44 |
| ≥85 | 32,065 (44.01) | 18,654 (31.30) | 0.26 | 45.45 | 0.03 | 44.01 |
| **Sex** |  |  |  |  |  |  |
| Male | 35,648 (48.93) | 27,083 (45.45) | 0.07 | 48.53 | 0.01 | 48.93 |
| Female | 37,204 (51.07) | 32,508 (54.55) | 0.07 | 51.47 | 0.01 | 51.07 |
| **Classification of department of residence** |  |  |  |  |  |  |
| Rural predominance | 21,912 (30.08) | 17,103 (28.70) | 0.03 | 30.04 | 0 | 30.08 |
| Urban predominance | 23,010 (31.58) | 20,466 (34.34) | 0.06 | 31.43 | 0 | 31.58 |
| Intermediate | 27,930 (38.34) | 22,022 (36.96) | 0.03 | 38.53 | 0 | 38.34 |
| **Deprivation index (FDep) quintiles**<br>(after imputation) |  |  |  |  |  |  |
| 1<sup>st</sup> quintile (least deprived) | 14,888 (20.44) | 11,835 (19.86) | 0.01 | 20.45 | 0 | 20.44 |
| 2<sup>nd</sup> quintile | 13,599 (18.67) | 10,518 (17.65) | 0.03 | 18.47 | 0 | 18.67 |
| 3<sup>rd</sup> quintile | 15,166 (20.82) | 12,126 (20.35) | 0.01 | 20.89 | 0 | 20.82 |
| 4<sup>th</sup> quintile | 14,688 (20.16) | 12,172 (20.43) | 0.01 | 20.17 | 0 | 20.16 |
| 5<sup>th</sup> quintile (most deprived) | 14,511 (19.92) | 12,940 (21.71) | 0.04 | 20.01 | 0 | 19.92 |
| **Type of residence** |  |  |  |  |  |  |
| Community dwelling | 64092 (87.98) | 56334 (94.53) | 0.23 | 86.79 | 0.04 | 87.98 |
| Nursing home / social residence | 8760 (12.02) | 3257 (5.47) | 0.23 | 13.21 | 0.04 | 12.02 |
| **Pre-existing chronic conditions** |  |  |  |  |  |  |
| None | 5537 (7.60) | 8405 (14.10) | 0.21 | 7.26 | 0.01 | 7.6 |
| One | 17,692 (24.28) | 16,471 (27.64) | 0.08 | 23.52 | 0.02 | 24.29 |
| Two | 23,257 (31.92) | 17,542 (29.44) | 0.05 | 31.76 | 0 | 31.92 |
| ≥ 3 | 26,366 (36.19) | 17,173 (28.82) | 0.16 | 37.46 | 0.03 | 36.19 |
| ***S. pneumoniae* vaccination** <sup>b</sup> | 12,238 (16.80) | 4732 (7.94) | 0.27 | 17.25 | 0.01 | 16.8 |
| **COVID-19 vaccination within 12 months before admission** |  |  |  |  |  |  |
| During season 2021-2022 | 6153 (8.45) | 3047 (5.11) | 0.13 | 8.33 | 0 | 8.45 |
| During season 2022-2023 | 8490 (11.65) | 4380 (7.35) | 0.15 | 11.76 | 0 | 11.65 |
| During season 2023-2024 | 5989 (8.22) | 1113 (1.87) | 0.29 | 9.07 | 0.03 | 8.22 |
| October to December 2024 | 388 (0.53) | 208 (0.35) | 0.03 | 0.58 | 0.01 | 0.53 |

Abbreviations: FDep: French deprivation index; SMD: standardized mean difference; IPTW: inverse probability of treatment weighting; EBAL: entropy balancing.
<sup>a</sup> Standardized mean difference between groups after entropy balancing is by default 0.00.
<sup>b</sup> Assessed during the 5 years before index admission.

During hospitalization, at least one acute cardiorespiratory, cerebrovascular, or renal complication was identified in 40% of patients (Supplementary Table-S4), occurring more frequently among those vaccinated than unvaccinated (42% vs 38.6%; p<0.001), reflecting the older age and greater burden of comorbidity among vaccinated patients before weighting. Co-diagnoses of bacterial or viral respiratory infections were identified in 7.6% vaccinated patients and 8.4% of unvaccinated patients (p<0.001). During the 2020–2024 seasons, SARS-CoV-2 accounted for 78% of documented viral co-infections.

### Association between influenza vaccination and disease outcomes

Across balancing strategies and COVID-19 vaccination adjustment scenarios, risk estimates were broadly consistent (Supplementary Table-S5). Because EBAL achieved better covariate balance and weight stability, EBAL-based models were retained for subsequent analyses. Among the approaches evaluated to account for the COVID-19 vaccination, models including interaction terms between COVID-19 vaccination and seasons were retained.

Compared with unvaccinated patients, vaccinated patients had a lower adjusted risk of IMV (aRR 0.88, 95%CI: 0.83 to 0.93), in-hospital all-cause death (aRR 0.90, 95%CI: 0.87 to 0.93), and the composite outcome of IMV, ICU admission, or death (aRR 0.96, 95%CI: 0.94 to 0.98) (Figure-1; Supplementary Table-S6). Vaccination was also associated with a lower crude risk of ICU admission (RR 0.95, 95%CI: 0.92 to 0.97), although this association was attenuated after adjustment (aRR 0.99, 95%CI: 0.96 to 1.01).

Associations were generally consistent across age groups (Figure-1; Supplementary Table-S7). Across epidemic seasons, the association between vaccination and lower risk of IMV was most consistently observed during the 2017–2018, 2018–2019, and 2019–2020 seasons (Figure-2; Supplementary Table-S8). Similarly, lower risk of in-hospital death among vaccinated patients was observed during the 2017–2018, 2018–2019, and 2022–2023 seasons.

**Figure 1.**
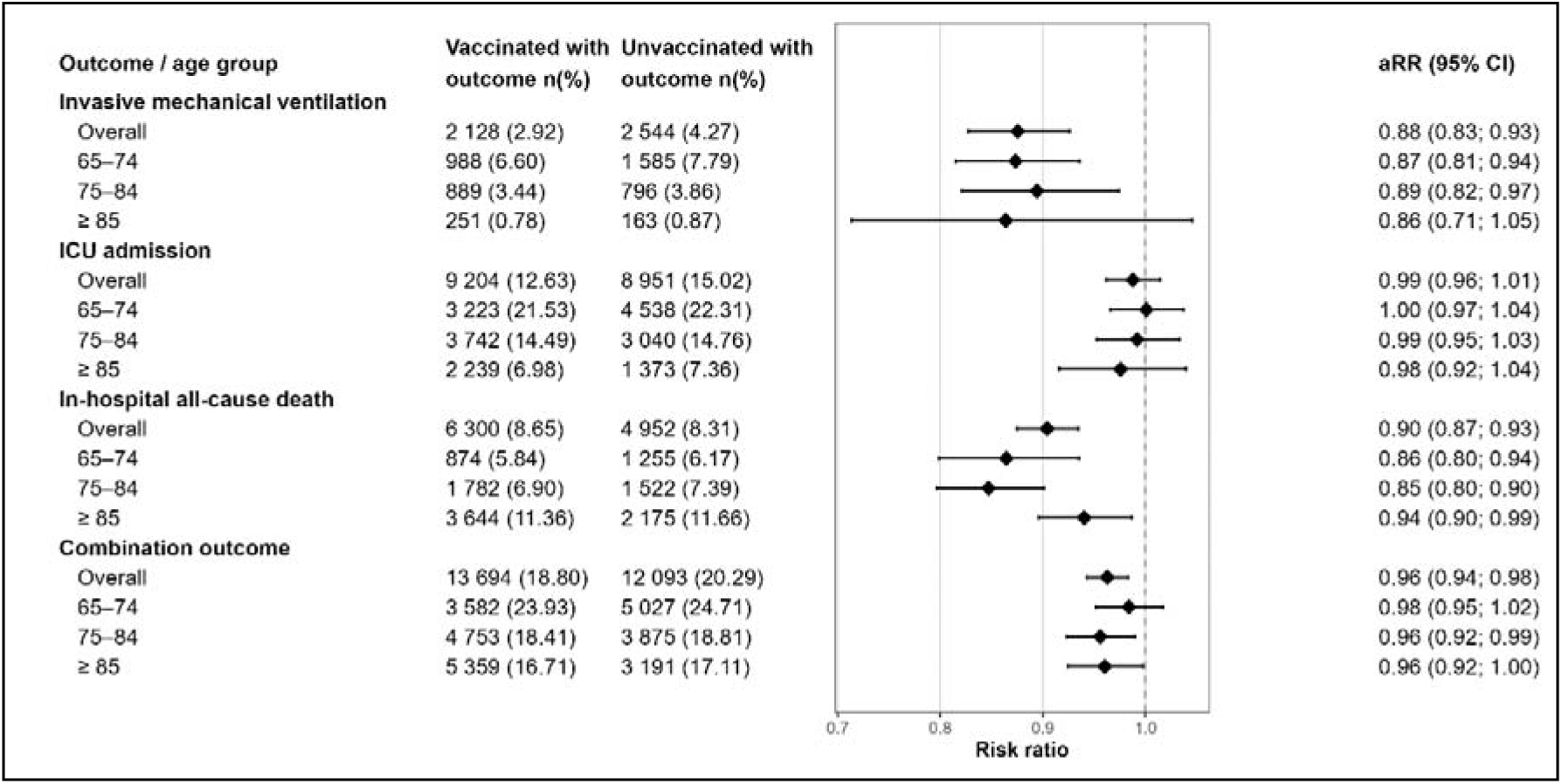
Association between vaccination status and main study outcomes after Entropy Balancing overall and by age group. Abbreviations: CI: Confidence interval; ICU: Intensive care unit; aRR: adjusted risk ratio. Combination outcome defined as need for invasive mechanical ventilation or intensive care unit admission or in-hospital death.

**Figure 2.**
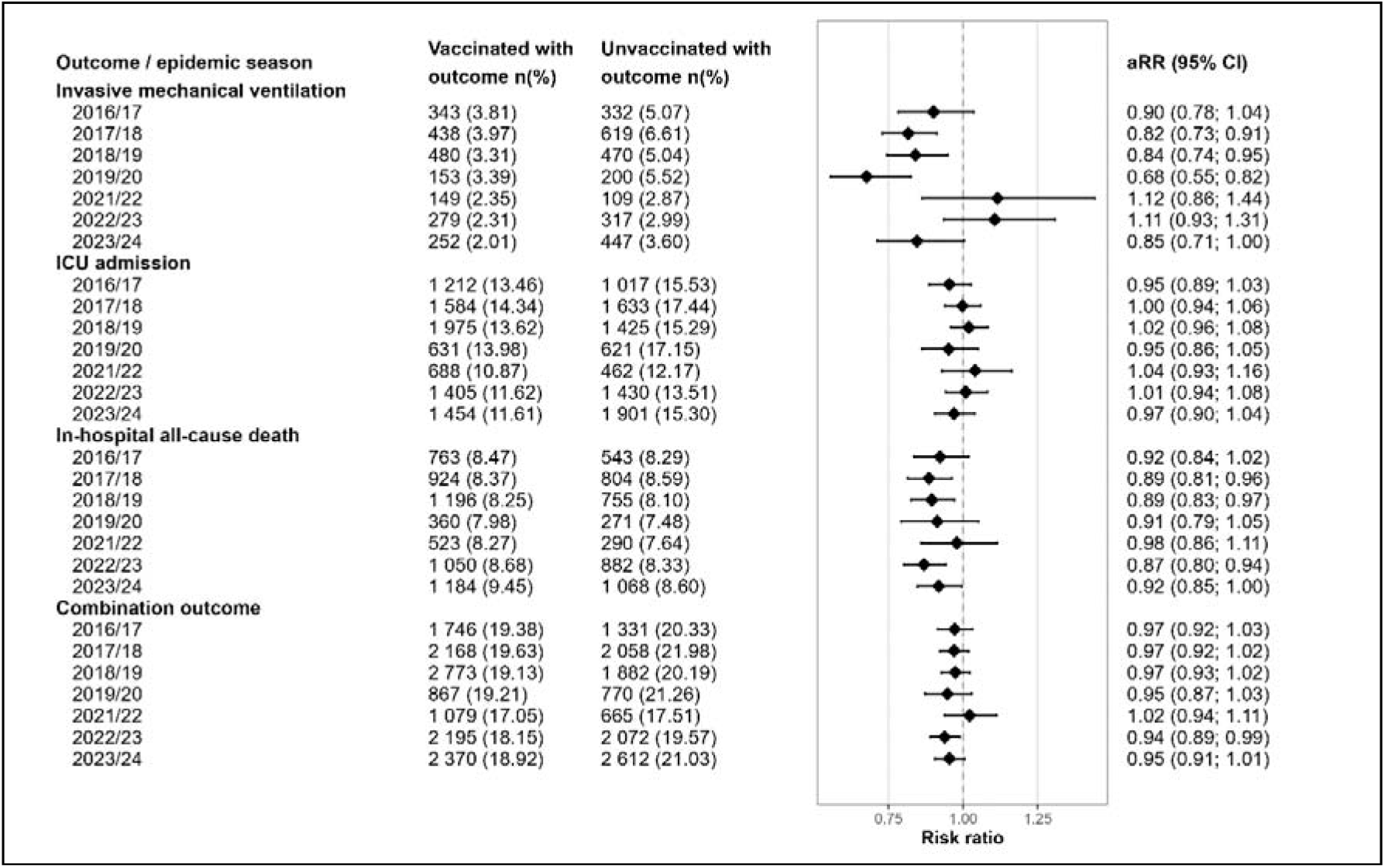
Association between vaccination status and main study outcomes after Entropy Balancing by season. Abbreviations: CI: Confidence interval; ICU: Intensive care unit; aRR: adjusted risk ratio. Combination outcome defined as need for invasive mechanical ventilation or intensive care unit admission or in-hospital death.

Analyses restricted to in-hospital deaths coded as influenza-related showed a similar direction of effect, although statistically significant reductions were observed only during the 2018– 2019 season, with cause-of-death information unavailable for the 2023–2024 season. During the season 2021-2022 vaccinated and unvaccinated patients had similar risks for all the main accessed outcomes.

Additional in-hospital and 30-day post-discharge outcomes are presented in Supplementary Table-S6. Vaccination was associated with lower risks of hospitalization lasting >5 days (aRR 0.97, 95%CI: 0.96 to 0.98) and admission to a reanimation unit (aRR 0.95, 95%CI: 0.91 to 0.99). The adjusted associations in terms of passive oxygen therapy and non-invasive mechanical ventilation did not reach statistical significance. Within 30-days after discharge, 6% (n=7220) of overall patients experienced rehospitalization for cardiorespiratory causes (aRR 0.98, 95%CI: 0.94 to 1.03) and 3.8 % (n=4554) died (aRR 0.88, 95%CI: 0.84 to 0.93).

### Multistate analyses of hospitalization trajectories

Multistate analyses were conducted to evaluate transitions over time between hospitalization, admission to ICU, in-hospital death and discharge alive within the first 30-days of hospital admission or within 30-days of ICU admission (Figure-3). Most ICU admissions and in-hospital deaths occurred during the first week of hospitalization, with divergence in post-ICU trajectories between vaccinated and unvaccinated patients becoming apparent shortly thereafter (Figure-3B). In balanced models, vaccinated and unvaccinated patients had similar hazards of transition to ICU over time (aHR 0.94, 95%CI: 0.88 to 1.00), aligned with the previous findings of absence of vaccination effect on ICU admissions, taking time into account and restricted to 30-days (Table-2). However, among patients admitted to ICU, vaccination was associated with a lower hazard of death (aHR 0.85, 95%CI: 0.78 to 0.92) whereas no effect of vaccination was observed on 30-day mortality in general hospitalization (aHR 0.96, 95%CI: 0.90 to 1.02) (Figure-3B, Table-2). Vaccinated patients had a higher probability of transition toward discharge both from general hospitalization (aHR 1.10, 95%CI: 1.09 to 1.12) or following ICU admission (aHR 1.10, 95%CI: 1.05 to 1.14) with 1-day and 2-days reduction in the median survival time to discharge, respectively (Figure-3).

**Table 2.**
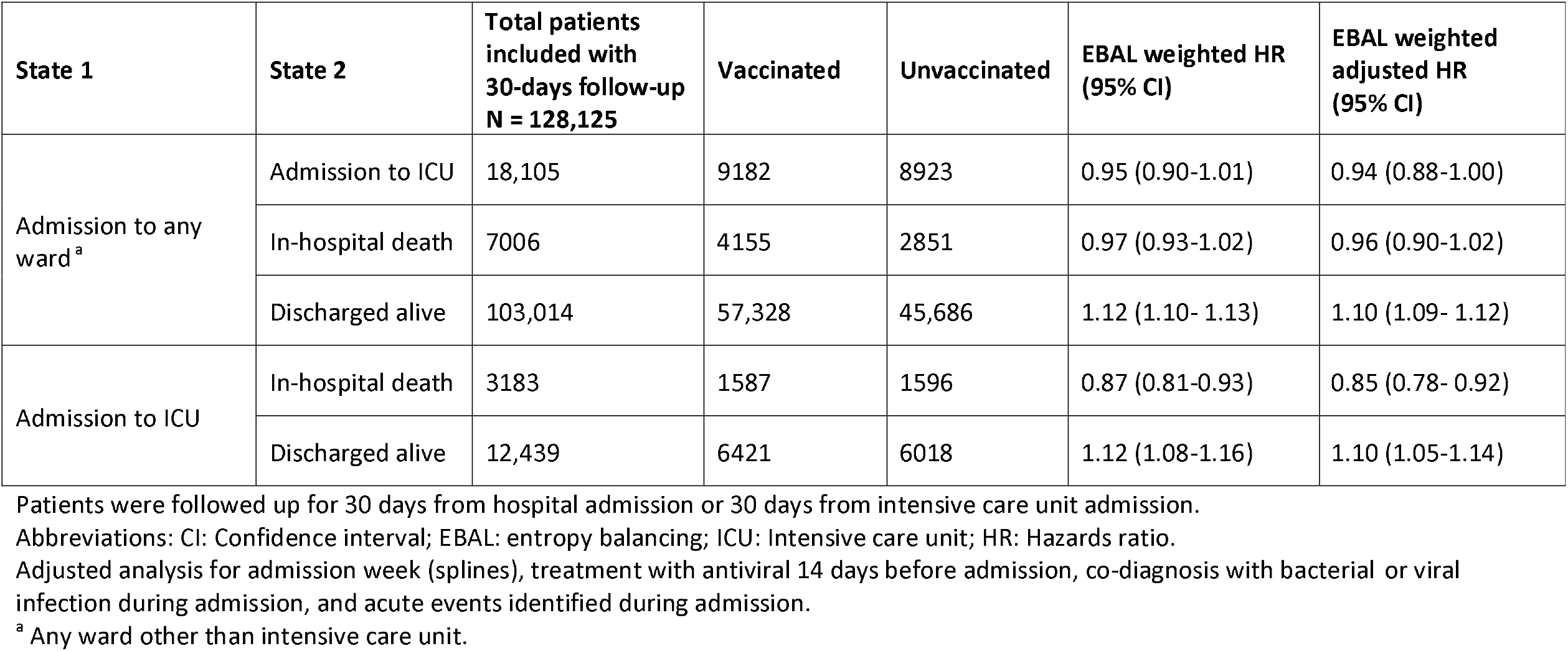
Multistate modelling of the association between vaccine status and in-hospital outcomes after Entropy Balancing.

**Figure 3.**
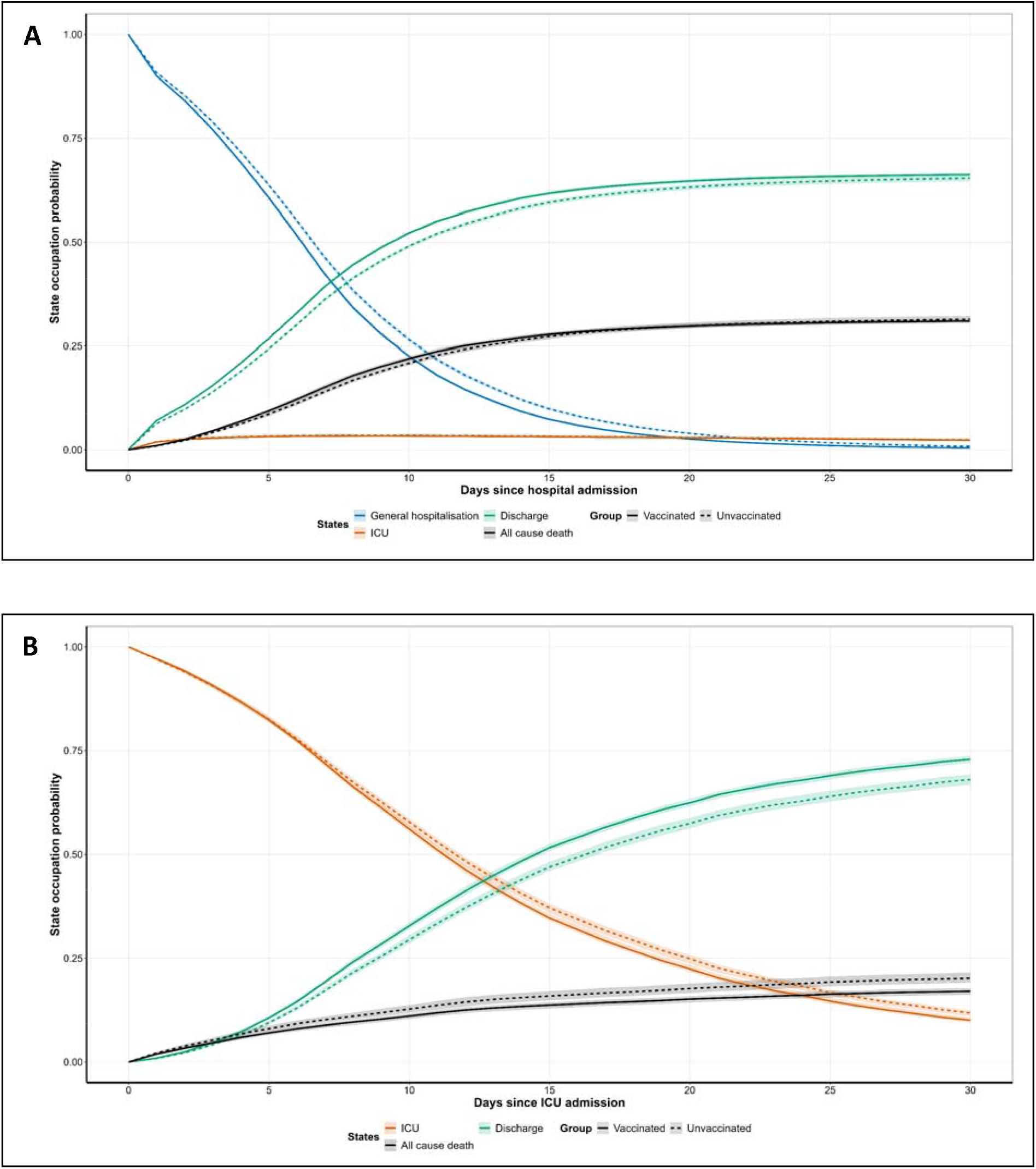
Multistate occupation probability curves in vaccinated and unvaccinated patients after Entropy Balancing **Panel A:** State occupation probability (and 95% confidence band) in the multistate model within 30 days of index admission in the vaccinated and the unvaccinated patients. The included states were admission to any unit, admission to intensive care unit, in-hospital all-cause death, and discharge alive. **Panel B:** State occupation probability (and 95% confidence band) within 30 days following admission to intensive care unit, including in-hospital all-cause death and discharge alive.

### Sensitivity analyses

Findings were generally consistent across sensitivity analyses (Supplementary Table-S9). Associations remained directionally consistent when analyses were restricting to hospitalizations with influenza as the primary diagnosis, including ICD-10 code J11, or excluding COVID-19 co-diagnosis. Associations were generally stronger when acute events identified during hospitalization were excluded from adjustment. Analyses restricted to patients with pneumonia as the primary diagnosis yielded similar but less precise estimates probably because of the smaller sample size.

## DISCUSSION

In this nationwide cohort study including more than 132,000 influenza-associated hospitalizations among adults aged ≥65 years across seven influenza seasons in France, influenza vaccination was associated with lower risks of invasive mechanical ventilation, in-hospital mortality, and a composite severe outcome among patients hospitalized with a diagnosis of influenza. Although the magnitude of these associations was modest, they were generally consistent across age groups and robust to multiple analytical approaches. Importantly, multistate analyses showed that vaccinated patients admitted to ICU experienced a lower risk of death and a faster transition towards discharge than unvaccinated patients, supporting that influenza vaccination may attenuate disease severity among older adults who experience breakthrough influenza hospitalization.

The role of influenza vaccination in attenuating disease severity among patients who develop influenza despite vaccination has received increasing attention. A review by Ferdinands et al. showed growing evidence supporting disease attenuation but noted substantial heterogeneity in outcome definitions, study designs, and populations, limiting comparisons across studies^12^. Earlier US studies also demonstrated that reductions varied by influenza season: during the 2012–2013 A(H3N2)-predominant season, vaccination showed only modest evidence of attenuation, whereas during the 2013–2014 season, when vaccine and circulating viruses were antigenically similar, vaccination among adults aged ≥65 years was associated with lower ICU admission (aOR 0.63), in-hospital death (aOR 0.39), and shorter hospital stay (aHR 1.24)^28,29^. In South America, vaccination among hospitalized adults aged ≥65 years was associated with lower in-hospital death (aOR 0.62), including death with and without ICU admission, but not with ICU admission itself or shorter stay^8^. In US claims data, high-dose vaccination was associated with lower 30-day post-influenza mortality compared with no vaccination across three seasons, with mortality reductions ranging 17%-29%, whereas standard-dose vaccination was associated with a significant reduction only in the 2016–2017 season (25%)^9^. More recently, a population-based cohort study in England found that influenza vaccination was associated with shorter hospital stays and lower mortality within six months after discharge among adults ≥65 years hospitalized with laboratory-confirmed influenza, but not with critical care admission or in-hospital death after adjustment^11^.

The reduction effects observed in our study were more modest than those previously reported. This difference should be interpreted in light of important methodological and programmatic differences. Unlike studies based on laboratory-confirmed influenza surveillance networks with detailed clinical information^8,11,28,29^, our study relied on nationwide administrative data and captured routine clinical practice across hospitals in France. This design maximizes population coverage and generalizability, but may also introduce greater heterogeneity in diagnostic coding, clinical management and thresholds for ICU admission. In addition, the vast majority of vaccinated patients in our cohort received standard-dose influenza vaccines, whereas enhanced vaccines have been associated with greater protection against severe outcomes among older adults^9,14^. Despite these differences, the direction of association across studies is consistent with a disease-attenuation effect of influenza vaccination, particularly for mortality among older adults hospitalized with influenza.

Vaccination was associated with a 12% lower risk of IMV and a 10% lower risk of in-hospital death. Although vaccination was not independently associated with ICU admission after adjustment, the multistate analyses suggested important differences in clinical trajectories following hospitalization. Among patients admitted to ICU, vaccination was associated with a 15% lower hazard of death and a higher probability of discharge alive. These findings suggest that vaccination status may be associated with outcomes occurring after progression to critical illness. Admission to ICU is also influenced by local clinical practice, resource availability, and thresholds for escalation of care, which may reduce its specificity as a marker of disease severity compared with mortality or IMV.

The effect of vaccination varied across epidemic seasons. Significant reductions in the risk of IMV were observed during the 2017–2018, 2018–2019 and 2019–2020 seasons, whereas lower in-hospital death was observed during the 2017–2018, 2018–2019 and 2022–2023 seasons. By contrast, no evidence of attenuation was observed during 2021–2022. Interpretation of this season requires consideration of its unusual epidemiological context. Following almost complete suppression of influenza circulation during 2020–2021, the 2021– 2022 epidemic in France was exceptionally late and characterized by altered transmission dynamics and co-circulation of multiple influenza strains^30^. Contemporary European studies also reported moderate vaccine effectiveness estimates during that season, particularly against A(H3N2) viruses^31^. In parallel, serological analyses suggested that the post-pandemic reduction in population protection was driven not only by reduced exposure to influenza viruses but also by the emergence of antigenically drifted strains^32^. Although our findings do not demonstrate a direct relationship between vaccine-virus matching and disease severity, they suggest that the magnitude of attenuation may depend partly on the broader virological and immunological context in which influenza epidemics occur.

To our knowledge, this is among the largest studies evaluating reduction of influenza severity among hospitalized older adults and one of the few using nationwide administrative data across multiple seasons. Our findings should therefore be interpreted in the context of both the strengths and limitations of real-world vaccine evaluation. Vaccinated patients were older and had a higher prevalence of chronic conditions than unvaccinated patients, reflecting current recommendations targeting individuals at increased risk of severe disease^33^. Such differences create substantial confounding by indication in observational studies of vaccine performance. To address this challenge, we compared two balancing approaches and several strategies to account for the COVID-19 vaccination era. Entropy balancing achieved high covariate balance and produced estimates comparable to those obtained using IPTW, supporting the robustness of the findings to the choice of weighting method. In addition, the near-complete coverage of the French population by the SNDS allowed evaluation of influenza-associated hospitalizations across public and private healthcare settings over multiple epidemic seasons.

Our results also have important public health implications. Despite longstanding vaccination recommendations and reimbursement policies in France, approximately half of influenza-associated hospitalizations in our study occurred among individuals without a recorded influenza vaccine dispensing during the corresponding season. Given the substantial burden of severe influenza among older adults, even relatively small reductions in severe outcomes may have meaningful population-level benefits. Moreover, substantial proportions of hospitalized patients experienced acute complications, and 6% required rehospitalization for cardiorespiratory causes within 30-days after discharge. Together, these findings reinforce the considerable clinical burden of influenza in ageing populations and support ongoing efforts to improve vaccine uptake while optimizing vaccination strategies for older adults^4^.

Several limitations should be considered. First, as with all studies based on administrative databases, information on smoking status, frailty, functional status, laboratory parameters, viral load, imaging findings, and treatments administered during hospitalization was unavailable. Residual confounding therefore remains possible despite adjustment and balancing procedures. Furthermore, restricting the analysis to hospitalized patients may introduce selection bias if vaccination influences the likelihood of hospitalization, potentially affecting associations between vaccination and subsequent disease severity. Second, influenza diagnoses were identified from ICD-10 codes and virological results were not available; therefore, some diagnostic misclassification is possible. Third, influenza vaccination status was inferred from reimbursed pharmacy dispensing records and may not perfectly reflect vaccine administration. To minimize exposure misclassification, we excluded residents of institutions with internal pharmacies where individual vaccine dispensing is not recorded in outpatient reimbursement databases. Nevertheless, some vaccinated individuals may have been misclassified as unvaccinated if vaccines were obtained through alternative mechanisms not captured in reimbursement records, although the magnitude of this bias is likely limited among community-dwelling older adults. Fourth, some acute respiratory, cardiovascular, cerebrovascular, and renal complications identified during hospitalization may lie on the causal pathway between influenza vaccination and severe outcomes. In sensitivity analyses excluding these variables from the adjustment set, associations between vaccination and severe outcomes were stronger, suggesting that adjustment for these events may have attenuated the estimated associations. Finally, cause-of-death coding in administrative databases is subject to misclassification and should therefore be interpreted cautiously. In addition, information on the medical cause of death was available only until the end of 2023.

In conclusion, influenza vaccination was associated with lower risks of IMV, in-hospital death, and mortality following ICU admission among older adults hospitalized with influenza in France. These findings provide population-level evidence consistent with attenuation of influenza disease severity among vaccinated patients requiring hospitalization, reinforcing the importance of improving vaccine uptake and optimizing vaccination strategies in older adults.

## Data Availability

The clinical codes, data management, and analysis codes used in this study are available on request from the corresponding author. Access to the data dictionary and these anonymised data can be granted upon request to the French National Health Data System platform.

## Contributors

CNAC designed the study, supervised the data management and analyses, and drafted the manuscript. ABD contributed to the data management and reviewed the manuscript. CNAC and ABD accessed the data at individual level. AS contributed to statistical analyses and reviewed the manuscript. BP contributed to the data management. HB and CES reviewed the manuscript. CC supervised the statistical analyses and reviewed the manuscript. MCN conceptualized the study, supervised the team, drafted the manuscript, accepts full responsibility for the work and the conduct of the study, and controlled the decision to publish. MCN attests that all listed authors meet authorship criteria and that no others meeting the criteria have been omitted.

## REFERENCES

1. World Health Organization. https://www.who.int/news-room/fact-sheets/detail/influenza-(seasonal)

2. European Centre for Disease Prevention and Control. Factsheet about Seasonal Influenza. https://www.ecdc.europa.eu/en/seasonal-influenza/facts/factsheet

3. Lafond KE, Porter RM, Whaley MJ, et al. Global burden of influenza-associated lower respiratory tract infections and hospitalizations among adults: A systematic review and meta-analysis. Kretzschmar MEE, ed. PLoS Med. 2021;18(3):e1003550. doi:10.1371/journal.pmed.1003550

4. Gavazzi G, Fougère B, Hanon O, et al. Enhanced influenza vaccination for older adults in Europe: a review of the current situation and expert recommendations for the future. Expert Review of Vaccines. 2025;24(1):350–364.

5. Langer J, Welch VL, Moran MM, et al. High Clinical Burden of Influenza Disease in Adults Aged ≥ 65 Years: Can We Do Better? A Systematic Literature Review. Adv Ther. 2023;40(4):1601–1627.

6. Bernard-Stoecklin S, Campèse C, Parent du Châtelet I. Fardeau de la grippe en France métropolitaine : bilan des données de surveillance des épidémies de 2011-2012 à 2021-2022. Published online 2023.

7. Santé publique France. Données Régionales de Couverture Vaccinale Grippe Par Saison et Dans Chaque Groupe d’âge. 2024. https://www.santepubliquefrance.fr/determinants-de-sante/vaccination/articles/donnees-regionales-de-couverture-vaccinale-grippe-par-saison-et-dans-chaque-groupe-d-age

8. Regan AK, Arriola CS, Couto P, et al. Severity of influenza illness by seasonal influenza vaccination status among hospitalised patients in four South American countries, 2013–19: a surveillance-based cohort study. Lancet Infect Dis. 2023;23(2):222–232.

9. Chaves SS, Naeger S, Lounaci K, et al. High-Dose Influenza Vaccine Is Associated With Reduced Mortality Among Older Adults With Breakthrough Influenza Even When There Is Poor Vaccine-Strain Match. Clin Infect Dis. 2023;77(7):1032–1042.

10. Presa J, Arranz-Herrero J, Alvarez-Losa L, et al. Influenza vaccine outcomes: a meta-analysis revealing morbidity benefits amid low infection prevention. Eur Respir Rev. 2025;34(175):240144.

11. Jeffery C, Cheyne CP, Buchan I, et al. Effect of influenza vaccination on post-admission outcomes for influenza patients in England: a population-based cohort study. Vaccine. 2025;68:127933. doi:10.1016/j.vaccine.2025.127933

12. Ferdinands JM, Thompson MG, Blanton L, Spencer S, Grant L, Fry AM. Does influenza vaccination attenuate the severity of breakthrough infections? A narrative review and recommendations for further research. Vaccine. 2021;39(28):3678–3695.

13. de Germay S, Conte C, Micallef J, et al. Performing pharmacoepidemiological studies using the French health insurance data warehouse (SNDS): How to translate guidelines into practice. Therapie. 2023;78(6):679–689.

14. Bricout H, Levant MC, Assi N, et al. The relative effectiveness of a high-dose quadrivalent influenza vaccine versus standard-dose quadrivalent influenza vaccines in older adults in France: a retrospective cohort study during the 2021-2022 influenza season. Clin Microbiol Infect. 2024;30(12):1592–1598.

15. Wyplosz B, Fernandes J, Sultan A, et al. Pneumococcal and influenza vaccination coverage among at-risk adults: A 5-year French national observational study. Vaccine. 2022;40(33):4911–4921.

16. Zysman M, Coquelin A, Le Guen N, et al. Prevalence and disparities in influenza vaccination among patients with COPD: A French nationwide population study. Respir Med. 2024;226:107606.

17. Tuppin P, Rudant J, Constantinou P, et al. Value of a national administrative database to guide public decisions: From the système national d’information interrégimes de l’Assurance Maladie (SNIIRAM) to the système national des données de santé (SNDS) in France. Rev Epidemiol Sante Publique. 2017;65 Suppl 4:S149–S167.

18. Équipes de surveillance de la grippe. Bulletin Épidémiologique Hebdomadaire. https://www.santepubliquefrance.fr/bulletin-epidemiologique-hebdomadaire

19. L’Institut national de la statistique et des études économiques. La France et Ses Territoires. 2021. https://www.insee.fr/fr/statistiques

20. Rey G, Jougla E, Fouillet A, Hémon D. Ecological association between a deprivation index and mortality in France over the period 1997-2001: variations with spatial scale, degree of urbanicity, age, gender and cause of death. BMC Public Health. 2009;9:33.

21. Rachas A, Gastaldi-Ménager C, Denis P, et al. The Economic Burden of Disease in France From the National Health Insurance Perspective: The Healthcare Expenditures and Conditions Mapping Used to Prepare the French Social Security Funding Act and the Public Health Act. Medical Care. 2022;60(9):655–664.

22. Van Buuren S, Groothuis-Oudshoorn K. mice: Multivariate imputation by chained equations in R. J Stat Softw. 2011;45:1–67.

23. Hainmueller J. Entropy Balancing for Causal Effects: A Multivariate Reweighting Method to Produce Balanced Samples in Observational Studies. Political Analysis. 2012;20(1):25–46.

24. Parish WJ, Keyes V, Beadles C, Kandilov A. Using entropy balancing to strengthen an observational cohort study design: lessons learned from an evaluation of a complex multi-state federal demonstration. Health Serv Outcomes Res Methodol. 2018;18(1):17–46.

25. Austin PC. An Introduction to Propensity Score Methods for Reducing the Effects of Confounding in Observational Studies. Multivariate Behav Res. 2011;46(3):399–424.

26. Localio AR, Margolis DJ, Berlin JA. Relative risks and confidence intervals were easily computed indirectly from multivariable logistic regression. J Clin Epidemiol. 2007;60(9):874–882.

27. Johansen R. An Empirical Transition Matrix for Non-homogeneous Markov Chains Based on Censored Observations. In: 1978. https://api.semanticscholar.org/CorpusID:124553225

28. Arriola CS, Anderson EJ, Baumbach J, et al. Does Influenza Vaccination Modify Influenza Severity? Data on Older Adults Hospitalized With Influenza During the 2012−2013 Season in the United States. J Infect Dis. 2015;212(8):1200–1208.

29. Arriola C, Garg S, Anderson EJ, et al. Influenza Vaccination Modifies Disease Severity Among Community-dwelling Adults Hospitalized With Influenza. Clini Infect Dis. 2017;65(8):1289–1297.

30. Équipes de surveillance de la grippe. Surveillance de La Grippe En France, Saison 2021-2022. 2022:362-375. http://beh.santepubliquefrance.fr/beh/2022/21/2022_21_1.html

31. Kissling E, Pozo F, Martínez-Baz I, et al. Influenza vaccine effectiveness against influenza A subtypes in Europe: Results from the 2021-2022 I-MOVE primary care multicentre study. Influenza Other Respir Viruses. 2023;17(1):e13069.

32. Fossum E, Rohringer A, Aune T, Rydland KM, Bragstad K, Hungnes O. Antigenic drift and immunity gap explain reduction in protective responses against influenza A(H1N1)pdm09 and A(H3N2) viruses during the COVID-19 pandemic: a cross-sectional study of human sera collected in 2019, 2021, 2022, and 2023. Virol J. 2024;21(1):57.

33. Assurance maladie. Campagnes de Vaccination. https://www.ameli.fr/assure/sante/assurance-maladie/campagnes-vaccination/vaccination-grippe

